# Sensitivity of RT-PCR testing of upper respiratory tract samples for SARS-CoV-2 in hospitalised patients: a retrospective cohort study

**DOI:** 10.1101/2020.06.19.20135756

**Authors:** Thomas C Williams, Elizabeth Wastnedge, Gina McAllister, Ramya Bhatia, Kate Cuschieri, Kallirroi Kefala, Fiona Hamilton, Ingólfur Johannessen, Ian F. Laurenson, Jill Shepherd, Alistair Stewart, Donald Waters, Helen Wise, Kate E. Templeton

**Affiliations:** MRC Human Genetics Unit, Institute of Genetics and Molecular Medicine, University of Edinburgh, UK; Clinical Microbiology & Virology, Directorate of Laboratory Medicine, Royal Infirmary of Edinburgh, NHS Lothian, Edinburgh, UK; Centre for Inflammation Research, University of Edinburgh, Edinburgh, UK; Edinburgh Critical Care Research Group, University of Edinburgh, Edinburgh, UK; eHealth Directorate, Royal Infirmary of Edinburgh, NHS Lothian; Blood Sciences, Directorate of Laboratory Medicine, Royal Infirmary of Edinburgh, NHS Lothian, Edinburgh, UK

**Keywords:** COVID-19, SARS-CoV-2, RT-PCR, Sensitivity and specificity, Diagnostics

## Abstract

**Objectives:** To determine the sensitivity and specificity of RT-PCR testing of upper respiratory tract (URT) samples from hospitalised patients with COVID-19, compared to the gold standard of a clinical diagnosis.

**Methods:** All URT RT-PCR testing for SARS-CoV-2 in NHS Lothian, Scotland, United Kingdom between the 7^th^ of February and 19^th^ April 2020 (inclusive) was reviewed, and hospitalised patients were identified. All URT RT-PCR tests were analysed for each patient to determine the sequence of negative and positive results. For those who were tested twice or more but never received a positive result, case records were reviewed, and a clinical diagnosis of COVID-19 allocated based on clinical features, discharge diagnosis, and radiology and haematology results. For those who had negative URT RT-PCR tests but a clinical diagnosis of COVID-19, respiratory samples were retested using a multiplex respiratory panel, a second SARS-CoV-2 RT-PCR assay, and a human RNase P control.

**Results:** Compared to the gold standard of a clinical diagnosis of COVID-19, the sensitivity of an initial URT RT-PCR for COVID-19 was 82.2% (95% confidence interval 79.0-85.1%). Two consecutive URT RT-PCR tests increased sensitivity to 90.6% (CI 88.0-92.7%). A further 2.2% and 0.9% of patients who received a clinical diagnosis of COVID-19 were positive on a third and fourth test.

**Conclusions:** The sensitivity of a single RT-PCR test of an URT sample in hospitalised patients is 82.2%. Sensitivity increases to 90.6% when patients are tested twice. A proportion of cases with clinically defined COVID-19 never test positive on URT RT-PCR despite repeated testing.

## Introduction

The Coronavirus disease 2019 (COVID-19) pandemic in Europe has already caused significant morbidity and mortality, not least within the United Kingdom. As well as causing large numbers of community acquired cases, SARS-CoV-2 has also been shown to circulate effectively within hospitals [1], necessitating the creation of COVID-19 specific areas. An estimate of the sensitivity of reverse transcription polymerase chain reaction (RT-PCR) testing for SARS-CoV-2 is therefore critical. Overestimation of sensitivity may lead to patients with disease being incorrectly diagnosed, and placed in non-COVID-19 areas with the subsequent risk of infection to others; underestimation of sensitivity may lead to patients who are SARS-CoV-2 negative being erroneously placed in COVID-19 areas.

A recent meta-analysis [2] estimates the sensitivity of RT-PCR testing of upper respiratory tract (URT) samples as 89%, but this meta-analysis, and a subsequent one [3] highlight a number of limitations in the literature. These include small sample size (<100 patients with COVID-19) [4–11], reliance on RT-PCR itself as the gold standard for diagnosis [12,13], use of computed tomography (CT) scans rather than clinical criteria as a gold standard for the diagnosis of COVID-19 [14,15], and absence of comprehensive RT-PCR testing for all included patients [16]. Finally, only a single study to our knowledge has examined the cumulative sensitivity of repeat testing for SARS-CoV-2 [14]. Here we examine in a large, comprehensive dataset the sensitivity of RT-PCR testing of upper respiratory tract specimens for COVID-19, compared to the gold standard of a clinical diagnosis.

## Methods

### Data source

All RT-PCR testing conducted for SARS-CoV-2 in NHS Lothian between the 7^th^ of February and 19^th^ April 2020 (inclusive) was reviewed. NHS Lothian covers a population of 907 580 people [17] and during the period of the study the Royal Infirmary of Edinburgh was the only regional centre conducting SARS-CoV-2 testing. Hospitalised patients were identified by cross-matching patient identification numbers against the NHS Lothian TrakCare Patient Clinical Management System database. In this study we comply with the STROBE reporting guidelines for observational studies[18].

### Data collection

For hospitalised patients, URT samples were identified, and only unambiguous positive or negative results, as authorised by laboratory staff, selected. Testing patterns were allocated for each patient, determining the sequence of URT RT-PCR tests and whether each test had yielded a negative or positive result (Table 1).

**Table 1.** Classification of test results.

| **Description** | **Classification** |
| --- | --- |
| Single negative test | Classified as a true negative. |
| Initial positive test, with or without subsequent testing. | Classified as a true positive. Clinical records reviewed to confirm that met clinical case criteria. |
| More than one negative test, no positive test result at any point | Clinical records reviewed to identify whether should be classified as true negative, or potential false negative based on clinical case criteria. |
| A series of one or more negative tests followed by a positive test, with or without subsequent testing. | Clinical records reviewed to identify whether a single or multiple clinical presentations. If two distinct clinical presentations with independent testing, treated as discrete episodes, and test classified as a true positive. If a single episode, negative test classified as a false negative. |

### Case definitions

Patients with a single negative test result were classified as a true negative, as clinical guidelines in place at the time specified that if there was clinical suspicion of COVID-19, an URT RT-PCR test should be repeated if the first test was negative. For those who initially tested negative on one or more occasions and then positive, case records were reviewed to determine whether this represented two discrete presentations or the same presentation. If they were classified as 2 distinct presentations, the negative followed by positive test was treated as a single positive test.

For those tested twice or more but who never received a positive result on URT RT-PCR testing, case records were reviewed, and a clinical diagnosis of COVID-19 was allocated based on a discharge diagnosis from the hospital team (or death certificate documentation) and further review of the case records. A positive clinical diagnosis was based on European Centres for Disease Control (ECDC) and World Health Organisation (WHO) criteria [19]. Based on previously published studies [20,21], cases were judged to be more likely to represent COVID if a chest XR showed patchy bilateral infiltrative changes, or a CT scan showed ground glass changes and if there was lymphopaenia in the presence of a normal neutrophil count [22]. Case records were reviewed by two clinicians (EW and TCW); if a consensus decision could not be reached, the case records were reviewed by a third clinician (DW) to arrive at a final clinical diagnosis. For patients classified as a possible false negative, their initial respiratory sample was retested using a multiplex respiratory panel, a second SARS-CoV-2 RT-PCR assay on the SeeGene platform as detailed below, and a human RNase P RT-PCR.

For patients who tested positive at any point, case records were reviewed to ensure they met the clinical case criteria for COVID-19, as described above. If they did not meet these clinical criteria, the samples IDs were matched against samples which had undergone whole genome sequencing (WGS) as part of the COVID-19 Genomics UK sequencing consortium [23]. If WGS had been completed successfully for a sample, this was assumed to represent a true positive. For those that had not, RT-PCR re-testing was conducted using the SeeGene platform as detailed below.

### Laboratory methods

URT samples were collected and added to viral transport media (Remel MicroTest M4RT). A volume of 110µL of eluate containing purified RNA was obtained following automated extraction carried out on the NucliSENS® easyMag® (bioMérieux) using an ‘off-board’ extraction where 200 µL of the sample was added to 2ml of easyMAG lysis buffer. The majority (93.7%) of tests on hospitalised patients were conducted using a modified in-house RT-PCR (Drosten, Eurosurveillance [24]); 6.3% were conducted using the Allplex™2019- nCoV Assay from SeeGene (Seoul, South Korea). The cut-off for diagnosis was a threshold cycle (Ct) of 40 or less.

### Further characterisation of possible false negatives

The Luminex Panel NxTAG® Respiratory Pathogen Panel (Texas, United States) was used to re-test the original extracted RNA for suspected false negatives (cases which met the clinical case criteria but had negative RT-PCR testing). Multiplex real-time PCRs were carried out on positive extracts using the ABI real-time system, model 7500 (Applied Biosystems, Warrington, United Kingdom), as part of routine testing using assays developed in-house and/or adapted from published methods [25,26]. The same samples were also re- tested using the Allplex™2019-nCoV SeeGene Assay, and using a human RNase P control [27]. For samples that tested positive using the SeeGene assay, Ct values for human RNase P were compared to negative results using a Welch two sample t-test in R [28] and plotted using GraphPad Prism version 6.04 for Windows (GraphPad Software, La Jolla California USA).

For patients who tested positive for a new respiratory pathogen, the case records were reviewed to ascertain whether the diagnosis was best explained by SARS-CoV-2 infection or the subsequently identified respiratory pathogen. Convalescent serology samples (>14 days after onset of symptoms), if available, were analysed using the Abbott SARS-CoV- 2 IgG assay on the Abbott Architect platform [29].

### Statistical analyses

The sensitivity was calculated as the proportion of true positives detected on initial testing and re-testing of suspected false negatives, divided by the number of true positives added to convincing false negatives, as estimated on the basis of further respiratory testing and serology testing. The specificity was calculated by dividing true negatives by the number of true negatives added to those judged to be false positives, on the basis of repeat RT-PCR retesting. The positive predictive value was determined by dividing the number of true positive by the sum of the true positives and false positives. The negative predictive value was calculated by dividing the number of true negatives by the sum of the true negatives and false negatives. Confidence intervals for these estimates were calculated using a two-sided exact binomial test with a confidence level of 0.95, implemented in R [28].

### Ethics statement

As part of the study protocol, specimens and associated clinical data were collected and anonymized before additional molecular /serological testing in accordance with local ethical approval (South East Scotland Scottish Academic Health Sciences Collaboration Human Annotated BioResource reference no. 10/S1402/33).

## Results

A total of 10601 RT-PCR tests for SARS-CoV2 for 8311 patients were conducted on URT specimens by the Royal Infirmary of Edinburgh laboratories between the 7^th^ of February and the 19^th^ of April 2020. These tests included community testing for patients who were never admitted to hospital, and testing for patients outside NHS Lothian for Boards that did not perform their own SARS-CoV2 testing. From this testing, 1667 patients received a positive result for SARS-CoV-2 testing (Table 2). The overall sensitivity of an initial URT RT-PCR test for the whole cohort (using a gold standard of an eventual molecular diagnosis of SARS-CoV-2 on URT RT-PCR) is 91.8%, rising to 98.4% after 2 tests.

**Table 2:** Summary of testing for all patients.

| Testing pattern | Number | % all patients | % of positive patients |
| --- | --- | --- | --- |
| Single negative test | 5665 | 68.1 | NA |
| More than 1 negative test | 979 | 11.8 | NA |
| Single positive test | 1531 | 18.4 | 91.8 |
| Initial negative test followed by positive test | 110 | 1.3 | 6.6 |
| Positive test after two or more negative tests | 26 | 0.3 | 1.6 |

### Testing for other respiratory pathogens

Of the total cohort 3226 patients were hospitalised within NHS Lothian, with 5418 tests. The data analysis for these patients is summarised in the flowchart in Figure 1. Seventy-three patients met the clinical case criteria for COVID-19 but did not receive a positive RT-PCR result (Figure 2). The RNA extract used for the initial SARS-CoV-2 RT-PCR was retested for common respiratory pathogens using the Luminex assay; 9 samples (12.3%) tested positive. On clinical review, all 9 cases were judged to be better explained by this new diagnosis rather than COVID-19 (Table 3).

**Table 3.**
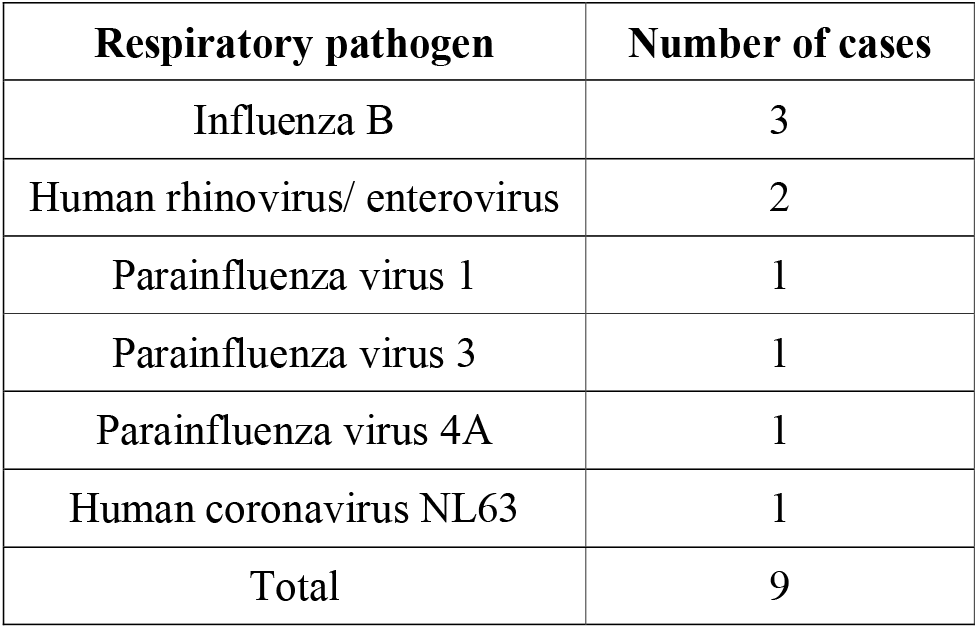
Positive results for other respiratory viruses on re-testing of initial sample.

| **Respiratory pathogen** | **Number of cases** |
| --- | --- |
| Influenza B | 3 |
| Human rhinovirus/ enterovirus | 2 |
| Parainfluenza virus 1 | 1 |
| Parainfluenza virus 3 | 1 |
| Parainfluenza virus 4A | 1 |
| Human coronavirus NL63 | 1 |
| Total | 9 |

**Figure 1.**
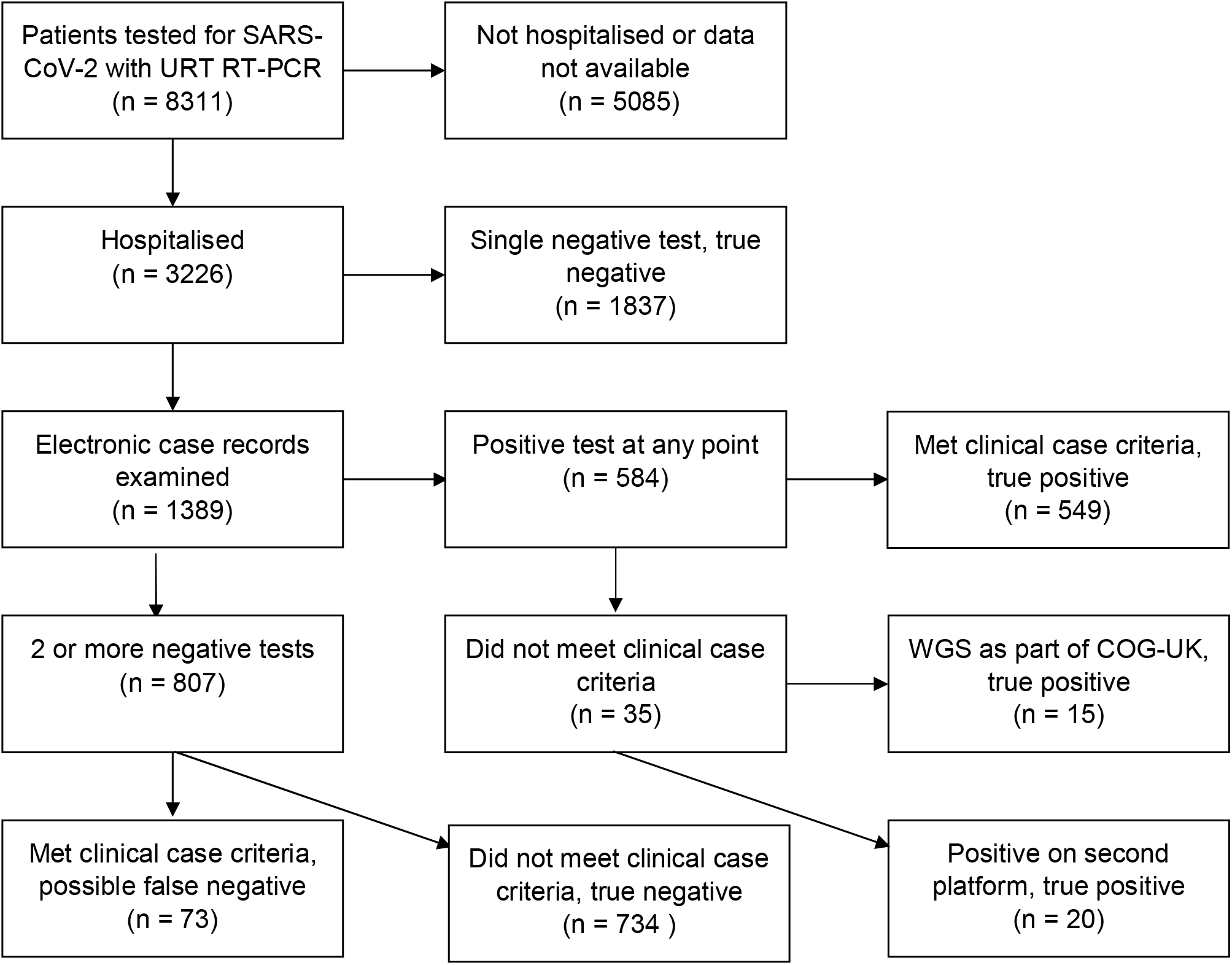
Flowchart for patients undergoing upper respiratory tract RT-PCR testing in NHS Lothian. URT: Upper Respiratory Tract. RT-PCR: Reverse transcription polymerase chain reaction. WGS: Whole genome sequencing. COG-UK: COVID19 Genomics Consortium.

**Figure 2.**
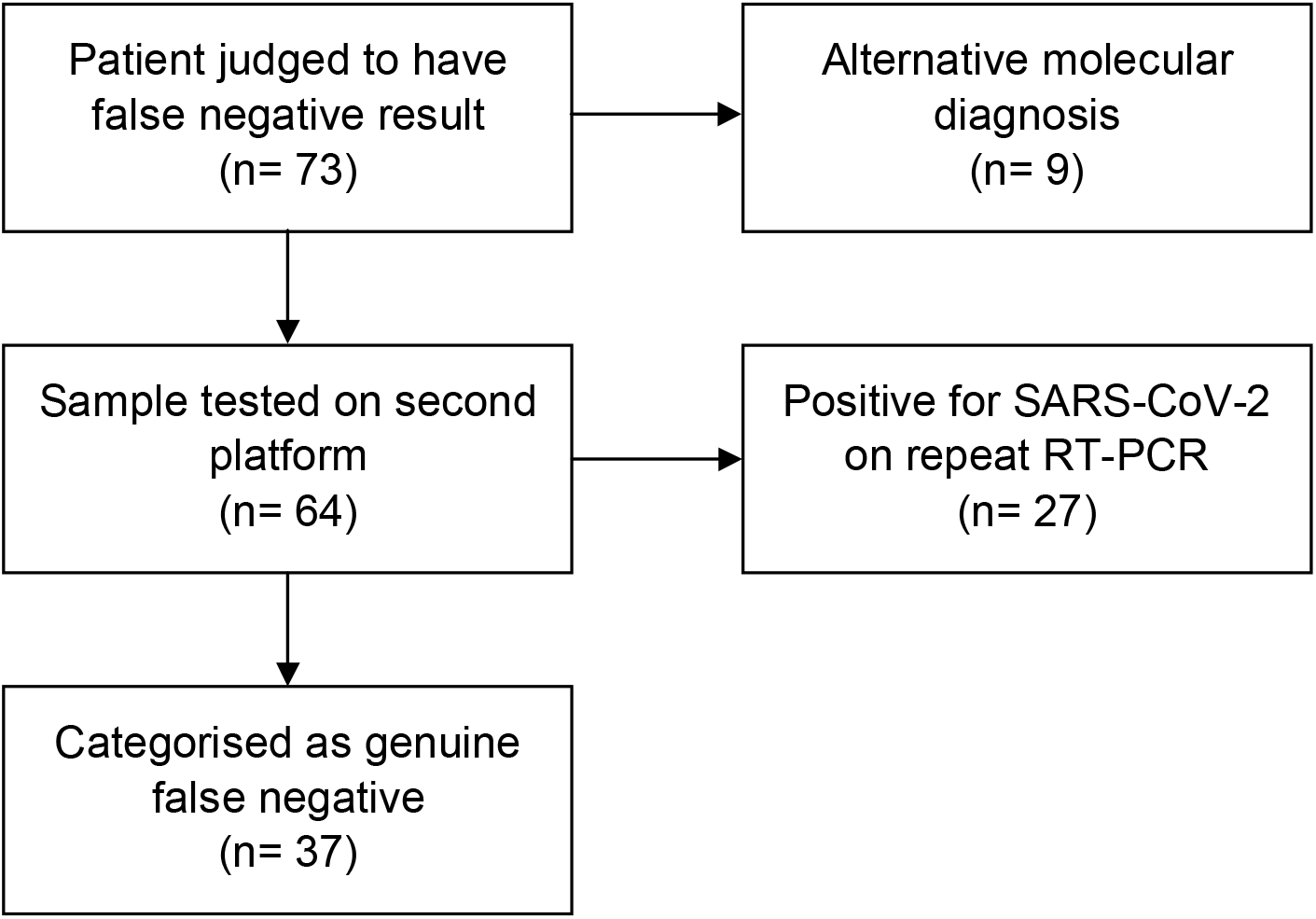
Flowchart for patients meeting clinical case criteria for COVID-19 but with negative upper respiratory tract RT-PCR testing.

### Retesting with the Seegene assay

All remaining 64 samples from suspected false negative cases had been tested initially with the modified in-house RT-PCR assay. Retesting with the Seegene assay for SARS-CoV-2 showed 27 (42.2%) of these were positive. Of the 37 remaining samples all showed a positive result for human RNase P. Comparing Ct values for human RNase P for SARS-CoV-2 positive and negative samples showed no significant difference using a Welch two sample t-test (p=0.49, Supplementary Figure 1).

### Sensitivity of initial test

For an initial test the sensitivity of RT-PCR URT for SARS-CoV-2 infection was 82.2% (95% confidence interval 79.0-85.1%) with a specificity of 100% (CI 99.9 – 100%). The positive predictive value of an initial test was 100%; the negative predictive value of an initial test was 95.7% (Table 4).

**Table 4.**
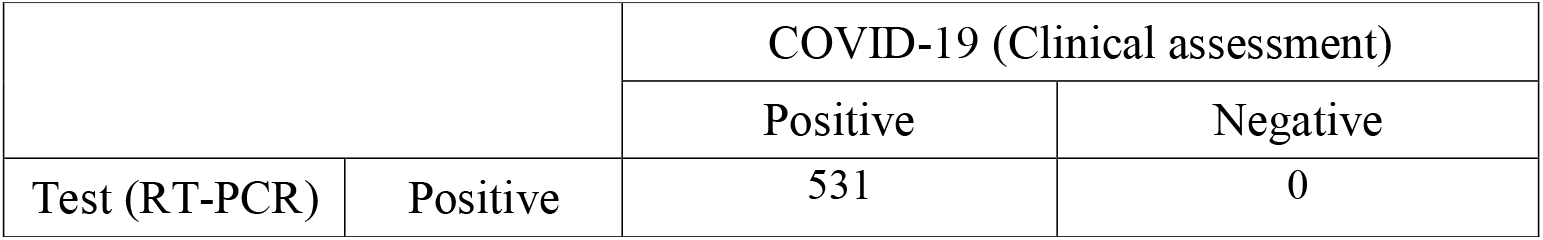
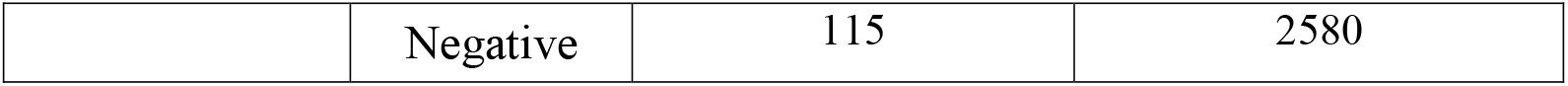
2 x 2 contingency table to calculate sensitivity and specificity of URT RT-PCR for SARS-CoV-2 detection on initial testing.

|  |  | COVID-19 (Clinical assessment) |  |
| --- | --- | --- | --- |
|  |  | Positive | Negative |
| Test (RT-PCR) | Positive | 531 | 0 |
|  | Negative | 115 | 2580 |

### Repeat testing

Sensitivity increased to 90.6 % (CI 88.0-92.7%) after 2 consecutive tests (Table 5), with a specificity of 100% (CI 99.9 – 100%). Increasing to 3 tests captured an additional 14/646 (2.2%) patients, and up to 4 tests an additional 6/646 (0.9%). This is a potential underestimate, as in this cohort there were 20 patients with a clinical diagnosis of COVID-19 who were tested twice with consecutive negative results, who might have yielded a positive result on a third test. The positive predictive value of 2 tests was 100%, and the negative predictive value 97.7%.

**Table 5.**
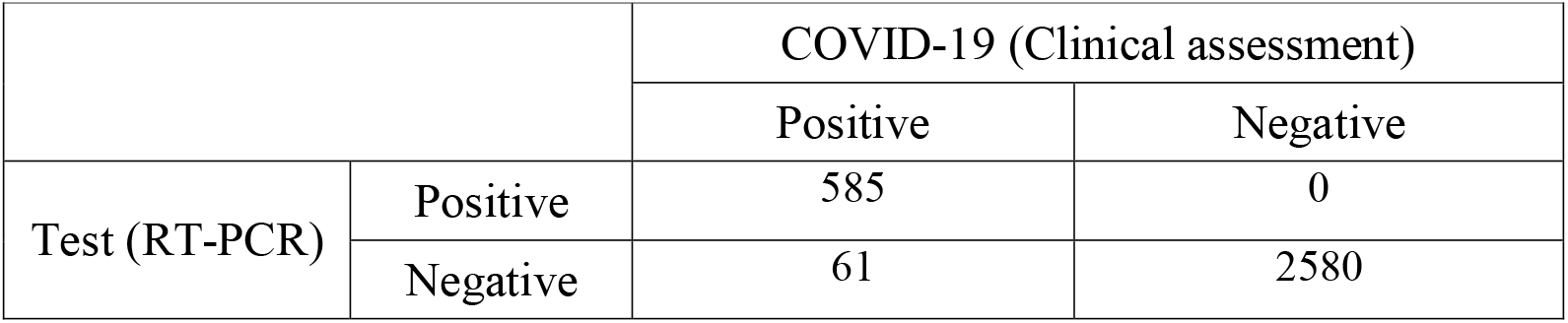
Sensitivity of 2 URT RT-PCR tests for the diagnosis of COVID-19.

|  |  | COVID-19 (Clinical assessment) |  |
| --- | --- | --- | --- |
|  |  | Positive | Negative |
| Test (RT-PCR) | Positive | 585 | 0 |
|  | Negative | 61 | 2580 |

### Lower respiratory tract samples

We examined data for a subset of 67 patients >16 years of age admitted to an Intensive Care Unit in NHS Lothian from the 6^th^ March until the 5^th^ of April 2020 with a discharge diagnosis of COVID-19. All tested positive on upper or lower respiratory tract RT-PCR testing. The sensitivity of an initial URT RT-PCR test in this cohort was 76.1% (51/67 positive, CI 64.1- 85.7%). After two URT RT-PCR tests, sensitivity increased to 89.5% (60/67 positive, CI 79.7-95.7%). Four patients never tested positive on URT RT-PCR (6.0%). Thirty-four patients had a lower respiratory tract (LRT) sample sent for RT-PCR: the sensitivity of this initial test was higher than that of URT testing at 94.1% (32/34 positive). This dataset, with the extra information offered by the availability of LRT specimens, supports the overall findings from the study.

### Convalescent serology

Out of the cohort of 64 patients who received a clinical diagnosis of COVID-19 with initial negative testing, and negative testing for other viruses, convalescent serology (>14 days after onset of symptoms) was available for 7 patients. Of these, 4 were positive (57.1%).

## Discussion

### Summary of principal findings

Here we show, using a comprehensively examined dataset, that the sensitivity of RT-PCR testing of URT specimens for the diagnosis of COVID-19 is 82.2% on initial testing, and 90.6% after two consecutive tests. Subsequent tests showed a small increase in diagnostic yield (2.2% for 3 tests and a further 0.9% for 4 tests), although this may represent an underestimate, as a number of patients given a diagnosis of COVID-19 based on clinical criteria were only tested twice.

### Findings of the present study in light of what has been published before

A previous meta-analysis gives a pooled sensitivity for RT-PCR of 89% (CI 81-94%) for the diagnosis of COVID-19 [2]; our results sit at the lower range of this estimate, but with overlapping confidence intervals. As highlighted in the introduction, the included studies suffer from a number of limitations including reliance on RT-PCR itself as the diagnostic gold standard, which would lead to an increase in the estimated sensitivity. We are not aware of any studies which have used a clinical diagnosis of COVID-19 against which to assess the sensitivity of RT-PCR. Here we show that the sensitivity of an initial test is lower than reported in this meta-analysis, but that the chance of a false negative result (17.8%) is lower than the 29% estimated in a subsequent meta-analysis [3] using a subset of studies included in [2]. These widely varying estimates highlight the importance of more data to inform our understanding of the strengths and weaknesses of RT-PCR testing.

### Strengths and limitations

The strengths of the study include the large dataset of both COVID-19 positive and negative patients, and extensive further testing to rule out false negative RT-PCR results and alternative diagnoses in those patients given a clinical diagnosis of COVID-19. We also studied whether suboptimal sampling might be a possible explanation for false negatives. However in a cohort of 37 possible false negatives all samples had detectable RT-PCR for human RNase P, with no difference between this group and those that tested positive for SARS-CoV-2, showing that this was not a factor in determining the sensitivity of RT-PCR in this population.

A limitation of the study is that the WHO/ECDC case definition of COVID-19 is likely to be highly sensitive but have low specificity. This means that a number of the cases we identified as potential false negatives could in fact represent other case presentations (a false positive in terms of the clinical diagnosis), and thus underestimate the sensitivity of the assay. This interpretation is supported by the findings from serology, where 4 out of 7 patients who met the clinical case criteria and had a convalescent serology sample had a positive serological test. Conversely, we did not examine the case records of the 1837 patients who tested negative on a single occasion, some of whom are likely to have received a clinical diagnosis of COVID-19. An increased number of false negatives would lead to a decreased sensitivity for the assay.

### Meaning of the study and understanding possible mechanisms

The result from our study suggest that there may be a small proportion of patients with SARS-CoV2 infection who meet the clinical case definition but never test positive on RT-PCR testing. It is possible that, in patients with severe disease, infection is entirely in the LRT, or that by time of presentation in the disease course the virus may only be present at very low levels in the URT [30]; this is supported by our findings in the ICU cohort, where 6.0% of patients never tested positive on URT RT-PCR.

### Implications for practice or policy, and suggestions for future research

Reliance on RT-PCR testing may result in patients with COVID-19 being inappropriately labelled with alternative diagnoses. These possibly infectious patients will subsequently pose a risk to healthcare workers and other patients. A more detailed picture of the sensitivity of URT RT-PCR testing will be aided by comprehensive serological testing of hospitalised patients with suspected infection.

## Data Availability

NA

## Acknowledgments

The authors would like to acknowledge the efforts of the clinical staff who collected samples from patients, and those of the laboratory staff at the Royal Infirmary of Edinburgh who processed them. They would also like to thank the members of the COG-UK team in Edinburgh who conducted the whole genome sequencing referenced in the study.

## ICMJE Statement

TCW, EW, GM and KET conceived and designed the study. AS, FH, KC and KK contributed towards the acquisition of data. GM, RB and HW conducted experimental work. TCW, EW, GM, KET, DW, IS and IFL conducted the analysis and interpretation of data. All the authors participated in drafting the article and revising it critically for important intellectual content, and all authors gave final approval of the version to be submitted.

## Figure captions

**Supplementary Figure 1:** Comparison of Ct values for human RNase P in URT RT-PCR tests which were positive or negative for SARS-CoV-2. Mean and standard deviation shown, p= 0.49 using Welch two sample t-test.

